# It is time to consider sleep in female athletes: assessment and tailored sleep intervention

**DOI:** 10.1101/2025.06.24.25330112

**Authors:** Mathias Goldberg, Benoit Pairot de Fontenay, Arnaud Boutin, Joffrey Cohn, Aurélie Sanjullian, Emeric Stauffer, Ursula Debarnot

## Abstract

**Purpose:** Optimal sleep is essential for supporting athletes’ health, performance and facilitating recovery. However, athletes’ sleep is impaired in both quality and quantity. Previous research has shown athletes’ sleep quality and quantity to be improved using sleep interventions, however very few studies have evaluated sleep and investigated it among female athletes. Therefore, this exploratory study aimed to evaluate the sleep of adult female team-sport athletes over a match week in an ecological context and to assess the effects of a one-to-one sleep intervention (comprising both educational and practical components).

**Methods:** Nine female rugby players completed a two-week protocol including a sleep evaluation (PRE week) and an intervention (POST week). The first week was dedicated to sleep assessment via both actigraphy and self-report questionnaires, and during the second week, players received educational and practical sleep interventions.

**Results:** During the PRE week, 88.9% of the participants were classified as poor sleepers, compared to only 44% during the POST week. A significant positive effect of sleep interventions was particularly found on subjective sleep (from 10.3 ± 1.1 to 8.8 ± 1.8, p = 0.03, η2 = 0.38) and psychological state (from 3.7 ± 0.6 to 2.9 ± 0.9, *p* = 0.002, *η^2^* = 0.55).

**Conclusion:** These positive effects highlight the need for sleep among female athletes and the potential benefits of targeted sleep interventions, though confirmation in larger samples of athletic women is needed. This feasible and tailored approach could be effectively implemented among female team-sport athletes over the competitive season.

## INTRODUCTION

Optimal sleep is essential for supporting athletes’ physiological health and psychological well-being, enhancing performance and facilitating recovery processes (Bird, 2013; Halson, 2014). Conversely, sleep deprivation or restriction has been reported to affect sport-specific skill execution, coordination and cognitive functions, mood, submaximal exercise or muscular and anaerobic power development (Bonnar et al., 2018; Fullagar et al., 2015; Walsh et al., 2021). Sleep disturbances concern 50-78 % of elite athletes (Walsh et al., 2021) and may be even more prevalent among recreational athletes, who must balance training with work, personal responsibilities and the pressures of an ‘always connected’ society, often without access to the logistical or professional support available to elites (Lucidi et al., 2007; Randell et al., 2021). In this context, sport-specific stressors such as high training loads, evening competitions, early sessions, and travel (Walsh et al., 2021) may have an even greater impact on their sleep. To date, sleep research in sport has mainly focused on males, leading to a significant underrepresentation of females. While emerging evidence suggests that females may be more vulnerable to extended wakefulness and circadian misalignment compared to males (Lok et al., 2024), sleep studies involving females have been conducted with clinical populations (e.g., obstructive sleep apnea, insomnia, restless legs syndrome and hypersomnia) or during specific life stages such as puberty, pregnancy or menopause (Lok et al., 2024).

Sleep must be evaluated by considering its multidimensional nature, incorporating both subjective perceptions and objective measurements (Bonnar et al., 2018). Considering the subjective dimension, initial studies in the general females population suggest that they tend to report lower perceived sleep quality compared to males using the Pittsburgh Sleep Quality Index (Fatima et al., 2016; Vitiello et al., 2004). In the context of sports science, similar trends have been observed using an athlete-specific, validated questionnaire, namely the Athlete Sleep Screening Questionnaire (ASSQ) (Xu & Li, 2024). Assessed by the sleep difficulty score (SDS) from the ASSQ, collegiate female swimmers experienced more sleep-related difficulties than males (8.0 ± 2.7 vs. 5.5 ± 1.5) (Xu & Li, 2024). The subjective dimension of sleep reflects individual self-perception and sex differences may partly be explained by psychological factors (Fatima et al., 2016). As females are twice as likely as males to develop anxiety disorders (Bandelow & Michaelis, 2015; Schaal et al., 2011), they may also be more vulnerable to sleep disturbances, with additional biological factors potentially contributing to this susceptibility (Fatima et al., 2016). Research involving female populations requires consideration of the menstrual cycle, as hormonal fluctuations and menstrual disorders can influence both sleep and circadian rhythms (Baker & Lee, 2022). Although the mechanisms by which ovarian hormones regulate sleep are not fully understood, sleep quantity has been shown to fluctuate across the cycle and is associated with the number of menstrual symptoms (Halson et al., 2024). Symptoms such as mental fatigue and pain are closely linked to sleep disturbances and may contribute to monthly sleep variations (Bixler et al., 2009; Halson et al., 2024). Reduced sleep quality, characterized by frequent awakenings, non-restorative sleep or nightmares (Lok et al., 2024; Miles et al., 2022), seems to occur in the days before or during the week of menstruation (Baker & Lee, 2022; Halson et al., 2024). Females tend to have an earlier chronotype than males (marked by an earlier peak in core body temperature and melatonin secretion), making sleep initiation more challenging due to evening training/competition and/or social/family demands (Lok et al., 2024). Considering these various subjective factors, it appears essential to evaluate sleep quality among female athletes while also considering objective sleep dimension.

In regard to objective sleep features, it has previously been reported that, compared to males, females have longer sleep quantity (total sleep time, TST, 6.0 ± 0.7 vs. 5.0 ± 0.4; Xu & Li, 2024), higher sleep efficiency (SE, 87.1 ± 5.2 vs. 76.7 ± 8.9; Xu & Li, 2024) and improved sleep architecture (higher per cent slow wave sleep, 5.0 ± 0.2 vs. 3.0 ± 0.3; lower percentage of light stage 1 sleep, 5.4 ± 0.2 vs. 14.1 ± 0.3; longer *Rapid-eye movement sleep* latency, 138.3 ± 3.0 vs. 104.9 ± 2.5; Bixler et al., 2009). The good objective sleep indices observed in females point to a discordance between objective and subjective sleep assessment (Gooderick et al., 2024; Vitiello et al., 2004). These observations emphasize the need to account for both subjective and objective dimensions of sleep, especially given the high sleep variability observed in female soccer players over a week (Costa et al., 2019). Together, these findings highlight specific sleep-related challenges in females, which are likely amplified in athletes, emphasizing the importance of developing interventions to address their individual needs.

Sleep interventions are based on traditional educational and practical approaches that have been widely used but have only recently been evaluated (Vachon et al., 2023; Pasquier et al., 2023; O’Donnell & Driller, 2017). In the context of female team-sport athletes, early work has objectively evaluated the effects of a single session of sleep hygiene education combined with relaxation strategies (e.g., progressive muscle relaxation, PMR) on elite netball athletes during pre-season, reporting an increase in TST as measured by actigraphy (O’Donnell & Driller, 2017). During their sleep education session, the focus was placed on the importance of sleep in athletes, sleep physiology, and general sleep hygiene, including five broad practical tips. However, these guidelines were not tailored to individual sleep needs, even though each athlete may require personalized recommendations (Bonnar et al., 2018), particularly in recreational sport settings where interventions must account for the specific life constraints of each player. Although subjective sleep seems to be the most affected dimension among females, they focused solely on objective sleep. To provide effective sleep interventions, assessment should integrate both objective and subjective measures over at least a week to account for night-to-night variability (Bonnar et al., 2018; Walsh et al., 2021). To our knowledge, no study has examined female athletes’ sleep using a multidimensional assessment over the competition phase while providing individually tailored interventions based on their specific sleep characteristics. Therefore, the present study aimed to evaluate the sleep of female adult team-sport athletes over a match week in an ecological context and assess the effects of a specific educational and practical sleep intervention. We hypothesized that females would exhibit altered daily subjective sleep quality and that a tailored sleep intervention could improve daily and weekly subjective sleep.

## MATERIALS AND METHODS

### Participants

Eleven females from a French recreational rugby union team were included in this two-week exploratory study. During the second week, two players were excluded because they were injured. Thus, nine players completed the two experimental weeks (22.3 ± 3.9 years, range: 18 – 28; height: 165.7 ± 6.4 cm; body mass: 77.1 ± 17.4 kg; BMI: 28.0 ± 5.4 kg·m-2; rugby roles of the players: six forwards and three backs; Table 1). Participants trained twice a week (Wednesday and Friday evenings at 8 p.m. for 1.5 hours) and had a match on Saturday at 5 p.m. Four players are working, three are full-time students, and two are enrolled in work-study programs. They gave written informed consent before taking part in the experiment. This study was conducted in accordance with current national and international laws and regulations governing the use of human subjects (Declaration of Helsinki). The study protocol was approved by the *Hospices Civil de Lyon*’s ethical committee (23-112).

**Table 1.** Demographic characteristics of the sample (n = 36).

|  | Mean ± SD | Min - Max |
| --- | --- | --- |
| Age (year) | 22.3 ± 3.9 | 18 - 28 |
| Height (cm) | 165.7 ± 6.4 | 156 - 175 |
| Body weight (kg) | 77.1 ± 17.4 | 57 - 110 |
| BMI (kg·m<sup>-2</sup>) | 28.0 ± 5.3 | 22.9 - 38.5 |
| Chronotype - MEQ (score) | 54.6 ± 7.1 | 43 - 63 |
| ESS (score) | 9.3 ± 3.0 | 4 - 15 |
| STAI - B (score) | 51.9 ± 4.9 | 46 - 59 |

BMI, body mass index; MEQ, Morningness-Eveningness Questionnaire; ESS, Epworth Sleepiness Scale; STAI - B, State-Trait Anxiety Inventory.

### Experimental design

Before the start of the experiment, participants were asked to complete an initial questionnaire to evaluate their chronotype (Morningness-Eveningness Questionnaire, MEQ ; Horne & Östberg, 1976), their sleepiness (Epworth Sleepiness Scale, ESS; Johns, 1991) and their anxiety (State-Trait Anxiety Inventory, STAI-B; Spielberger, 1983). Participants were evaluated for two weeks (PRE / POST) separated by one month dedicated to processing data from the PRE week (Figure 1). During the two-week protocol, they wore an actimeter (GTX3+, ActiGraph, Pensacola, USA) on the non-dominant wrist 24 hours a day, except during showers and contact training sessions, to enable objective sleep assessment. This device has been validated against polysomnography (Quante et al., 2018), and actigraphy is recommended for monitoring sleep in team-sport players (Bonnar et al., 2018; Cunha et al., 2023). Actilife LLC Pro software v6.13.4 (Actigraph, Pensacola, USA) was used to analyze wake-sleep cycle and sleep data with the Cole-Kripe algorithm (60-second epoch sample). Actimeter provides the following variables: 1) bedtime (h:min), 2) wake-up time (h:min), 3) total sleep time (min), 4) sleep efficiency (%), 5) wake after sleep onset (min), and 6) coefficient of variation of TST (CV_TST_).

**Figure 1.**
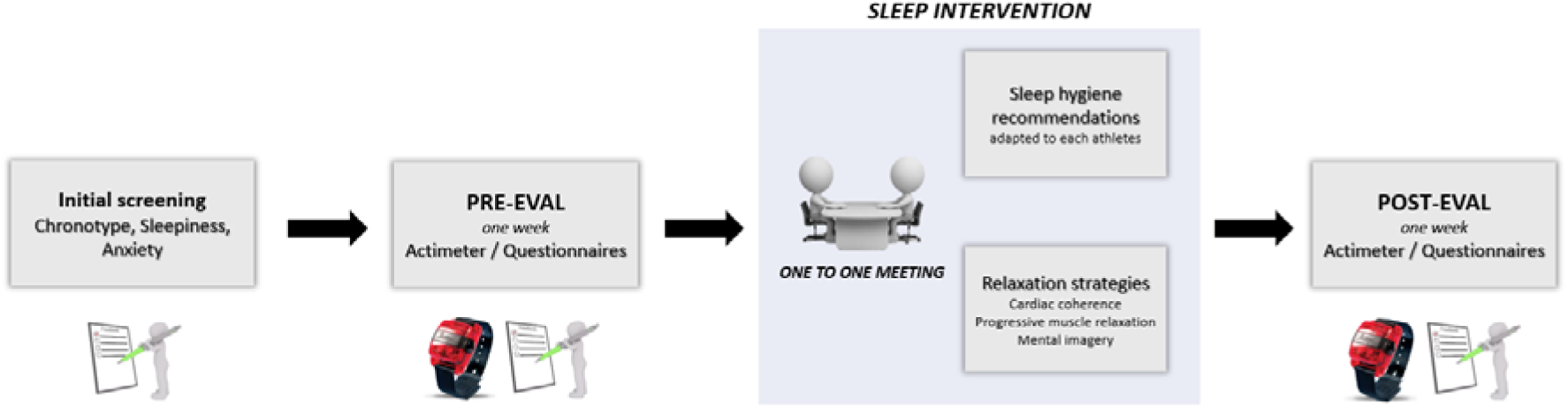
Experimental design.

At the beginning of the PRE week and the end of the POST week, participants completed the ASSQ, which provided a weekly subjective SDS. Participants also answered a modified version of the Hooper and Mackinnon questionnaire (1995) (Hooper_m_) (Hooper & Mackinnon, 1995) daily during the two weeks (PRE/POST) within the first hour after waking up. This questionnaire assessed fatigue (1 – 7 scale; ‘how fatigued are you?’), sleep quality (1 – 6 scale; ‘how was your sleep last night?’), sleep quantity (1 – 7 scale; ‘how many hours did you sleep last night?’), muscle soreness (1 – 7 scale; ‘rate your level of muscle soreness?’), and psychological state (1 – 7 scale; how are you feeling psychologically?’). Additional item assessing sleep latency (1 – 6 scale; ‘how long it took you to fall asleep?’) was completed to estimate time of sleep onset. Based on the Hooper_m_ questionnaire, we summed scores on sleep quality, quantity, and latency to measure daily subjective sleep. In this study, a total score of ≤ 9 out of 19 was as indicative of good subjective sleep quality.

To combine objective and subjective sleep dimensions, we used a Sleep composite score that we have previously developed considering SDS, daily subjective sleep, SE and CV_TST_ (Goldberg et al., *preprint* 2025) and calculated as follows :

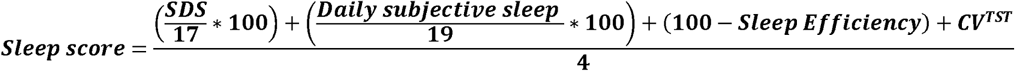

This score was calculated based on the literature (SDS ≤ 4, daily subjective sleep ≤ 9, SE ≥ 85%). In our previous study, we have decided to consider CV_TST_ = 19.5 %, as cut-off value, based on the prior findings of Saidi et al. (2023) among young rugby players. However, as sleep differs between males and females, we have decided to consider in this study the previous result from Costa et al. (2019) for the CV_TST_ (= 15.2) among female soccer players. Taking together, participants needed a sleep composite score lower than 25.4 to be considered good sleepers. Moreover, participants reported their daily screen time by estimating total usage across devices (e.g., phone, TV, computer, tablet), and provided a daily indication of menstrual symptoms by answering the question: ‘Over the past day and night, did you experience any menstrual cycle-related pain (e.g., abdominal pain)?’ (YES/NO).

Sleep intervention was provided the Sunday before the POST week. Participants attended a one-to-one session (30’) with the same sleep expert (M.G., PhD candidate in Neuroscience and sports physiotherapist specializing in sleep and recovery strategies), during which they were educated on the physiological and psychological impacts of sleep, with a specific focus on athletic performance, recovery and injury risk. Based on the sleep-related issues identified during the PRE week, participants received individualized sleep hygiene recommendations, which they were instructed to implement daily over the POST week. For example, players with an early chronotype but a delayed bedtime were educated in this misalignment and encouraged to advance their bedtime. Those presenting with sleep debt were advised to extend their nocturnal sleep duration, incorporate daytime naps, and go to bed earlier. Participants who reported difficulties initiating sleep were guided in establishing a consistent sleep routine, including avoiding heavy meals and screen exposure (at least 1 h) before bedtime, practicing relaxation techniques, and maintaining regular sleep-wake. These educational components were adapted, combined and tailored to address the specific needs and sleep profiles of each athlete. Finally, M.G. introduced three relaxation strategies, namely cardiac coherence, progressive muscle relaxation, and mental imagery, highlighting their recognized benefits for stress regulation and sleep (e.g., deeper sleep, reduced sleep onset latency, and enhanced relaxation). Following the educational session, participants were provided with audio recordings on their phone (5’ each ; Jacobson, 1938; Perreault-Pierre, 2012; McCraty & Zayas, 2014; Walker, 2017), and were encouraged to use them as guided practices when needed. After discussing the feasibility and integration of the recommendations into their daily routines, participants were encouraged to implement at least two during the POST week. They left the session with a personalized leaflet summarizing their individual sleep recommendations. During the POST week, participants indicated daily whether they had implemented a recommendation and, if so, specified which one.

### Statistical analysis

The primary outcome was the daily subjective sleep. Secondary outcomes were objective sleep variables (bedtime, wake-up time, TST, SE, WASO and coefficient of variation of TST), other subjective sleep variables (SDS, Hooper_m_, daily screen time) and the sleep composite score. Using established cut-off values in line with current recommendations, the number of participants meeting the criteria for TST (i.e., > 7 hours), SE (i.e., > 85%) and WASO (i.e., < 20 minutes) were analyzed (Ohayon et al., 2017). As residuals followed linearity, homoscedasticity and normality (Shapiro-Wilk test), linear mixed models were implemented to assess the effect of the intervention between PRE and POST weeks. The significant threshold (*p*) was set at 0.05 for all analysis. Effect sizes (*η^2^*) were calculated to interpret the magnitude of the mean difference between the two weeks or days of the week/weekends (small [0.01], medium [0.06] and large [0.14]; Cohen, 1988). Statistical analysis was performed using R software (version i386 4.1.2). As proposed by Ditroilo et al. (2025), a sensitivity power analysis was conducted a posteriori using G*Power (v. 3.1.9.7) to determine the minimum detectable effect size with the current sample size (Lakens, 2022).

## RESULTS

Based on the initial questionnaires, all players exhibited either intermediate (five players) or moderate morning (four players) chronotypes. Regarding the ESS, two players showed no signs of sleepiness, six exhibited signs of sleepiness, and one showed signs of excessive sleepiness. Additionally, seven players had moderate anxiety levels, while two players demonstrated high anxiety levels (STAI). PRE and POST sleep assessments are presented in Table 2.

**Table 2.** Objective and subjective sleep data throughout the night in PRE and POST weeks (mean and ± SD).

| INTERVENTION | PRE | POST | *p* | *N*<sup>2</sup> |
| --- | --- | --- | --- | --- |
| **Subjective variables** |  |  |  |  |
| ASSQ – SDS (score) | 5.4 ± 2.5 | 4.6 ± 2.4 | **0.0006\*\*\*** | 0.59 |
| Sleep quality (score) | 3.5 ± 0.9 | 2.8 ± 0.7 | **0.04\*** | 0.33 |
| Sleep quantity (score) | 4.5 ± 0.8 | 4.0 ± 1.0 | 0.15 | 0.20 |
| Sleep latency (score) | 2.4 ± 0.5 | 1.9 ± 0.6 | 0.9 | 0.26 |
| Daily subjective sleep (score) | 10.3 ± 1.1 | 8.8 ± 1.8 | **0.03\*** | 0.38 |
| Fatigue (score) | 4.0 ± 0.9 | 3.2 ± 1.4 | **0.03\*** | 0.37 |
| Muscle soreness (score) | 2.5 ± 1.0 | 2.0 ± 1.1 | 0.08 | 0.28 |
| Psychological state (score) | 3.7 ± 0.6 | 2.9 ± 0.9 | **0.002\*\*** | 0.55 |
| Hooper<sub>m</sub> (score) | 20.5 ± 1.8 | 16.8 ± 3.7 | **0.006\*\*** | 0.49 |
| Screen time exposure (min) | 04:32 ± 02:07 | 04:34 ± 02:18 | 0.9 | 0.001 |
| **Objectives variables** |  |  |  |  |
| Bedtime (hh:mm) | 00:11 ± 00:59 | 00:08 ± 01:34 | 0.9 | 0.002 |
| Wake-up time (hh:mm) | 07:56 ± 00:44 | 07:51 ± 00:40 | 0.6 | 0.03 |
| Sleep efficiency (%) | 88.6 ± 8.0 | 90.3 ± 3.3 | 0.4 | 0.06 |
| Total sleep time (min) | 410.7 ± 51.3 | 439.8 ± 51.9 | 0.1 | 0.22 |
| CV<sub>TST</sub> (%) | 22.4 ± 12.5 | 17.5 ± 9.4 | 0.3 | 0.10 |
| Wake after sleep onset (min) | 53.4 ± 43.5 | 45.9 ± 18.0 | 0.5 | 0.05 |
| Sleep composite score | 30.0 ± 4.6 | 25.0 ± 7.3 | 0.06 | 0.30 |

CV<sub>TST</sub>, Coefficient of variation of total sleep time. ASSQ – SDS, Athlete Sleep Screening Questionnaire – Sleep Difficulty Score. The effects between PRE and POST weeks are indicated by “\*” ( $p \leq 0.05$ ), “\*\*” ( $p \leq 0.01$ ) and “\*\*\*” ( $p \leq 0.001$ ). For score variables, a low score means good sleep quality. Sleep quality (1 – 6 scale), sleep quantity (1 – 7 scale), sleep latency (1 – 6 scale), fatigue (1 – 7 scale), muscle soreness (1 – 7 scale), and psychological state (1 – 7 scale), Hooper<sub>m</sub> (6 – 40 scale).

A sensitivity power analysis, based on a repeated-measures design with two time points (PRE and POST), nine participants, an alpha level of 0.05, and an assumed correlation of 0.5 between measurements, revealed that the study had 80 % power to detect an effect size of f = 0.53 (equivalent to *η²* = 0.22, a medium-to-large effect). All observed effects presented thereafter exceeded this threshold, indicating that the study was adequately powered to detect the observed effects. Regarding the subjective dimension, only one player reported good sleep quality (≤ 9) during PRE week, whereas five players met this threshold during the POST week, based on their daily subjective sleep scores. A significant positive effect of sleep interventions was found on daily subjective sleep (*p* = 0.03, *η^2^* = 0.38), SDS (*p* = 0.0006, *η^2^* = 0.59), Hooper (*p* = 0.006, *η^2^* = 0.49), and especially on sleep quality (*p* = 0.04, *η^2^* = 0.33), fatigue (*p* = 0.03, *η^2^* = 0.37) and psychological state (*p* = 0.002, *η^2^* = 0.55). In the objective dimension, TST was 410.7 ± 51.3 and 439.8 ± 51.9 minutes during PRE and POST weeks and CV_TST_ was 22.4 ± 12.5 and 17.5 ± 9.4, respectively. Compared to current sleep recommendations, five players met the > 7 h TST criterion during PRE and seven during POST; eight and nine players had SE > 85 % in PRE and POST respectively; no player had WASO < 20 minutes during PRE, and one did during POST. However, no statistical differences were found between the PRE and POST weeks among objective variables, nor for any other quantitative sleep variables. Combining both subjective and objective dimensions, the sleep composite score categorized eight players as bad sleepers during PRE week (88.9 %) while only four during POST week (44.4 %), suggesting a trend toward improvement in the sleep between the PRE and POST weeks (*p* = 0.06, *η^2^* = 0.30).

Regarding menstrual symptoms, 10 out of 126 recorded nights (∼ 8 %) were reported as being affected by menstrual cycle-related pain; however, no associations were found between these symptoms and sleep-related variables. Among the proposed sleep interventions, seven out of nine players adhered to their individualized recommendations during the POST week. Specifically, six players limited screen exposure in the hour before bedtime, six took naps, five extended their sleep duration, five practiced relaxation techniques, and four advanced their bedtime. Each recommendation could be implemented multiple times during the week by each player.

## DISCUSSION

The present study aimed to evaluate the sleep of female team-sport athletes and the effects of individualized sleep intervention. Our main findings revealed that (i) 88.9 % of the female players suffered sleep disturbances, with impairments observed in both subjective and objective sleep dimensions, and (ii) a single sleep intervention led to sleep subjective quality improvements. These findings provide novel and converging evidence supporting the importance of sleep assessment among female athletes and highlight the potential benefits of tailored interventions, aligning with emerging research in female athletic populations.

In our study, nearly all recreational female players experienced sleep disturbances across both objective and subjective measures, a prevalence even higher than previously reported in studies of elite male and female athletes (Goldberg et al., 2025; Lastella et al., 2015; O’Donnell & Driller, 2017; Vachon et al., 2023; Walsh et al., 2021). To the best of our knowledge, this is the first study to assess sleep in recreational female athletes using both objective and subjective assessments over a competition week. Regarding the subjective sleep dimension, players exhibited poor sleep quality at both the daily level (i.e., daily subjective sleep scores) and the weekly level (i.e., SDS scores). Notably, although the SDS scores in this study were lower, indicating better perceived sleep, than those previously reported in female swimmers (5.4 ± 2.5 vs 8.0 ± 2.7; Xu & Li 2024), they still exceed the threshold for sleep disturbances. Xu & Li (2024) also highlighted female athletes reported poorer subjective sleep quality than their male counterparts. More specifically, the moderate to high anxiety levels observed in all players, as reflected in STAI scores, may partly account for the reported subjective sleep disturbances, potentially through mechanisms such as pre-sleep ruminations, nightmares or early morning awakenings (Richards et al., 2020; Schaal et al., 2011). Objectively, players averaged less than 7 h of sleep per night, which is one hour less than the current recommendations for athletes (Walsh et al., 2021) and lower than the 7.8 hours observed among female athletes (Miles et al., 2022). This insufficient sleep quantity has been reported to hinder performance or recovery by reducing sleep-dependent restorative processes, such as the release of growth hormones essential for tissue repair (Fullagar et al., 2015). As expected, we further observed a high intraindividual variability in the sleep quantity (CV_TST_) and bedtime routine during the PRE week (Miles et al., 2022). This lack of sleep and high variability at the individual level may be attributed to the dual demands faced by recreational athletes, who must manage professional and lifestyle constraints, resulting in time pressure and the need to adapt their routines to optimize sports performance.

After sleep evaluation, an individualized intervention combining theoretical and practical relaxation strategies can lead to meaningful improvements in both daily and weekly subjective sleep quality. Given that females tend to report more impaired perceptions of sleep than males (Xu & Li, 2024), the observed improvements in subjective sleep quality provide new evidence supporting the integration of targeted sleep interventions into athletic training programs. Among the sleep disturbances identified in athletes, anxiety, pre-competition pressure, or higher mental workload are well-known phenomena that can result in a non-restorative sleep feeling (Miles et al., 2022; Schaal et al., 2011). To mitigate these psychological impairments, seven of the nine players adhered to their individualized recommendations. Among the two remaining players, one reported minimal sleep disturbances throughout the study, while the other was unable to adopt new sleep behaviors due to professional constraints, including frequent on-call duties and multiple jobs. This indicates that all players for whom the recommendations were both relevant and feasible successfully implemented them, likely motivated by the awareness raised during the one-to-one session regarding their importance and practicality. Among the others, most adopted pre-bedtime and morning routines, such as limiting screen time exposure before sleep, practicing relaxation strategies, extending sleep duration, and going to bed earlier. Although a reduction in screen exposure during the POST week was expected and players reported limiting their use in the hour before bedtime, no significant change was observed between PRE and POST assessments. This discrepancy may be explained by a shift in screen use to earlier of the day or to the complexity of the question, which asked participants to estimate total screen time across multiple devices such as phones, TV, computers, and tablets. This may have introduced inaccuracies in screen time self-reported data.

Among objective sleep dimensions, all variables showed a trend toward improvement over the POST week, although these changes did not reach statistical significance. While previous studies have reported increases in TST after sleep interventions (O’Donnell & Driller, 2017), such improvements were not significant in the present study, likely due to high interindividual variability among athletes and the limited sample size, despite the changes approaching sensitivity thresholds. In addition, objective sleep changes may require more time to emerge, particularly given the lifestyle constraints faced by recreational athletes compared to professionals (O’Donnell & Driller, 2017). This likely contributes to the absence of significant changes in objective sleep measures observed in this one-week intervention. Interestingly, the study by O’Donnell and Driller (2017) suggests that for teams without access to a sleep expert or facing time constraints for individualized education, group-based sleep education may represent a valuable first step toward improving sleep quantity. Interestingly, when combining both objective and subjective sleep dimensions, the sleep composite score revealed a trend toward a global sleep improvement in the current study. These findings suggest that this individual sleep intervention influenced both dimensions in a similar direction and may offer be implemented with the aim to promote all female athletes’ sleep indices.

### Practical implications

According to a recent paper on recovery strategies, while coaches appear to be well aware of the importance of sleep, athletes tend to place less emphasis on it (Li et al., 2024). Sleep is an invisible part of training and is therefore sometimes ignored, unlike an injury that immediately alerts athletes and limits their participation in sports. As observed in this study, raising sleep awareness seems to have a significant and effective impact on female team sport athletes. To begin, it is crucial to establish a sleep profile for each athlete, considering tools such as the MEQ, ESS, and ASSQ, along with the Athlete Sleep Behaviour Questionnaire (Driller et al., 2018), to accurately identify sleep habits. Coaches should then work with sleep experts to assess both objective and subjective sleep dimensions over at least one week (Bonnar et al., 2018; Walsh et al., 2021). This duration allows for the weekly variations assessment and helps tailor specific sleep interventions more effectively. Designed to change sleep behaviors, these interventions can be concise (30 minutes) and practical, incorporating both theoretical and practical components. To achieve optimal outcomes, sleep interventions should be tailored to each athlete’s specific needs and capacity to integrate them into their daily routines, especially in recreational athletes. Promoting adherence requires helping athletes understand the value of these strategies so they are not perceived as unnecessary or time-consuming. Notably, most athletes in this study chose to continue incorporating the sleep recommendations after the intervention week, having experienced positive effects in their daily lives.

### Limitations

Even if this original paper reports an interesting view of female team-sport athletes’ sleep, some limitations should be noted. First, although the cohort sample is small, the results observed here must be taken with precautions as exploratory study (Ditroilo et al., 2025), and further ecological studies must be conducted with female athletes to replicate and extend our findings. Secondly, sleep was not assessed with a polysomnographic device, thus not allowing a precise evaluation of the sleep architecture. As reported by Lok et al. (2024), females have a specific organization of NREM and REM stages during sleep cycles. Thus, it could be interesting to evaluate the effect of sleep interventions on team-sport females’ sleep architecture. Thirdly, this study did not objectively assess hormonal variations by collecting urine or blood samples. While hormonal variations over the menstrual cycle are well documented, assessing their evolution in relation to post-intervention sleep improvements (Baker & Lee, 2018) is an interesting perspective for future research.

## CONCLUSION

The present study provides a novel and early investigation of female athletes’ sleep by combining objective and subjective assessments over a competition week. Findings revealed insufficient sleep across both dimensions, which may negatively affect athletic performance and recovery, as well as broader physiological and psychological processes associated with poor sleep. Individualized sleep interventions led to improvements, particularly in the subjective sleep dimension. These encouraging findings warrant confirmation in future studies with appropriate error control. As females are more prone to subjective sleep disturbances, this type of rapid, feasible, and tailored assessment and intervention may represent an effective approach for improving sleep among female team-sport athletes.

## Data Availability

All data produced in the present study are available upon reasonable request to the authors.

## Declarations

### Funding or Competing interests

The authors have no competing interests to declare that are relevant to the content of this article. Ethics approval: The study protocol was approved by the *Hospices Civil de Lyon*’s ethical committee (23-112).

